# Inter-surgeon variation in reoperation following strabismus surgery among Medicare beneficiaries: Associations with adjustable sutures, patient and surgeon characteristics

**DOI:** 10.1101/2023.05.03.23289451

**Authors:** Christopher T. Leffler, Alicia Woock, Meagan Shinbashi, Melissa Suggs

## Abstract

**Purpose:** The objective of this study was to quantify inter-surgeon variation in strabismus surgery reoperation rates in a large national database of provider payments, and to explore associations of reoperation rate with practice type and volume, surgical techniques, and characteristics of the patient population.

**Methods:** Fee-for-service payments to providers for Medicare beneficiaries having strabismus surgery between 2012 and 2020 were retrospectively analyzed to identify reoperations in the same calendar year. The adjustable-suture technique was considered to be available to the patient if the patient’s surgeon billed for adjustable sutures. Predictors of the rate of reoperation for each surgeon were determined by multivariable linear regression.

**Results:** Among 141 surgeons, the reoperation rate for 1-horizontal muscle surgery varied between 0.0% and 30.8%. Due to the presence of high-volume surgeons with high reoperation rates, just 11 surgeons contributed half of the reoperation events for 1-horizontal muscle surgery in this national database. Use of adjustable sutures, surgeon gender, and surgical volume were not independently associated with surgeon reoperation rate. Associations of reoperation with patient characteristics, such as age and poverty, were explored. In a multivariable model, surgeons in the South tended to have a higher reoperation rate (p=0.03). Still, the multivariable model could explain only 16.3% of the variation in surgeon reoperation rate for 1-horizontal muscle. For 1-vertical muscle surgery, patient poverty was associated with a lower surgeon reoperation rate (p=0.008).

**Conclusions:** Patient-level analyses which ignore inter-surgeon variation will be dominated by the practices of a small number of high-volume, high-reoperation surgeons. There are order-of-magnitude variations in reoperation rates among strabismus surgeons, the cause of which remains largely unexplained.

## Introduction

Variation between surgeons in patient outcomes has been explored with respect to non-ophthalmic procedures,^1^ as well as ophthalmic procedures, such as cataract extraction^2^ and corneal transplantation.^3^ We sought to study variation between surgeons in outcomes from strabismus surgery. One commonly used outcome metric for strabismus surgery is reoperation rate.^4^ We sought to determine if variation between surgeons in strabismus surgery reoperation rate could be explained: 1) by characteristics of the surgical approach, such as the use of adjustable sutures; 2) by surgeon characteristics, such as gender, seniority, or practice volume; or 3) by aspects of the patient population in the practice, such as age or poverty.

We evaluated reoperations for strabismus surgery in the database of Medicare payments to providers for 2012 through 2020.^5^

## Methods

This study was approved by the hospital Office of Research Subjects Protection. We downloaded the national database of Medicare payments for 2012 through 2020.^6^ This database includes payment data for every practitioner in the country who received Medicare fee-for-service payments. In the United States, Medicare is a single-payer, national health insurance program administered by the federal government serving patients over age 65 and younger patients with disabilities. Each current procedural terminology (CPT) code had to be paid to the provider for at least 11 beneficiaries in 1 year for that particular CPT to be listed under that provider for the year. We also downloaded characteristics of patient clinical and demographic information for each provider’s Medicare patients, for the mid-point year (2016), or, if no data were available for this year for a particular provider, whichever year was closest to the mid-point year.^7^

We defined junior surgeons as those who entered the Medicare database during the 2012 to 2020 period, senior surgeons as those who left the Medicare database during this period, and remaining surgeons as mid-career.

We evaluated rates of reimbursed reoperations in patients having one horizontal muscle strabismus surgery (CPT 67311), one vertical muscle surgery (CPT 67314), adjustable suture placement (CPT 67335), and surgery with scarring of extraocular muscles (e.g., prior ocular injury, strabismus, or retinal detachment surgery) or restrictive myopathy (e.g., dysthyroid ophthalmopathy; CPT 67332). These CPT codes were selected because they are the most commonly coded strabismus procedures.

Other strabismus surgery codes were not used frequently enough to draw meaningful conclusions. The reoperation rate for each surgeon was determined from the numbers of beneficiaries and beneficiary service days. For instance, if a given provider treated 13 beneficiaries with a particular CPT code in a given year, but there were 14 beneficiary service days for this code, then 1 of the 13 beneficiaries had a reoperation. The unit of analysis was the surgeon. If the surgeon received any payments for CPT 67335, then the adjustable-suture technique was considered to be available to the provider.

We compared the likelihood of reoperation in patients having strabismus surgery when the adjustable technique was available with patients having surgery when the adjustable technique was not available. We also evaluated associations of reoperation rate with academic or community-based practice, and surgery in a practice with the lowest or highest surgical volume by quartile. Reoperation rate was evaluated in major geographic regions – Northeast (CT, ME, MA, NH, RI, VT, NJ, NY, PA), Midwest (IN, IL, MI, OH, WI, IA, KA, MN, MO, NE, ND, SD), South (DE, MD, DC, FL, GA, NC, SC, VA, WV, AL, KY, MS, TN, AR, LA, OK, TX), and West (AZ, CO, ID, NM, MT, UT, NV, WY, AK, CA, HI, OR, WA).^8^ We excluded data from retinal oncologists, who might have been coding for muscle surgeries when detaching muscles to place radiotherapy plaques.

Reoperation rates were compared by the t-test. Practices were grouped according to median population values. Significant variables in univariate analysis were analyzed with multivariable linear regression. Availability of adjustable sutures was included in the model because of the clinical interest in this question.

## Results

### Horizontal Muscle Surgery

Among 141 surgeons coding for 1-horizontal muscle surgery (CPT 67311), the average reoperation rate was 4.93% (SD 5.77%), with a median value of 3.57%.

However, a wide range was observed, from 0 to 30.8% (Figure 1).

**Figure 1.**
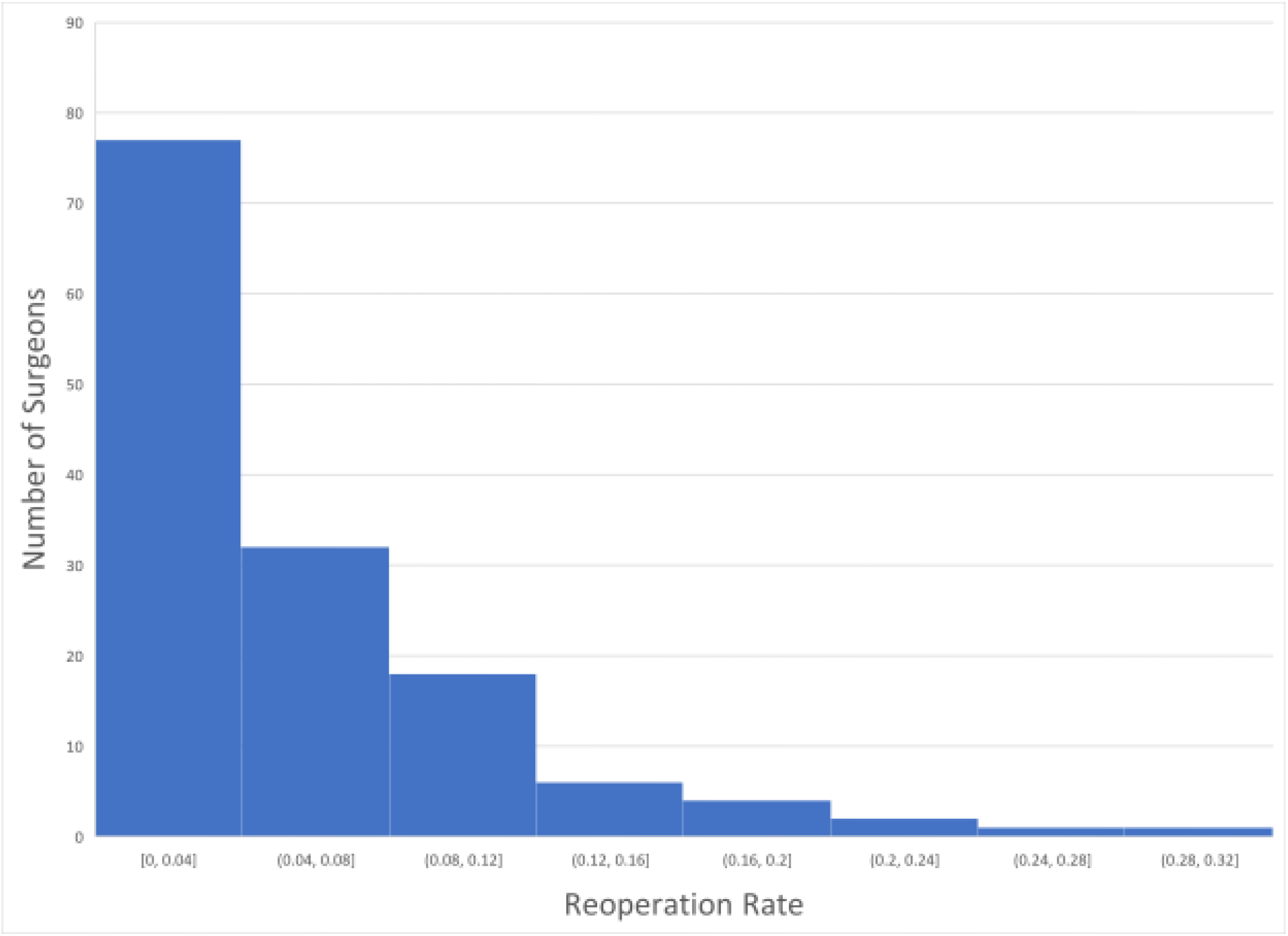
Histogram of reoperation rate among 141 surgeons for one-horizontal muscle surgery (CPT 67311).

Average patient age of at least 71 years in the practice was associated with a higher reoperation rate (5.68% vs. 3.65%, p=0.03, Table 2). In addition, prevalence of qualification for Medicaid (a marker of poverty) at or above the median value (15.17%) was associated with a lower reoperation rate (3.71% vs. 6.33%, p=0.01, Table 2).

**Table 1.**
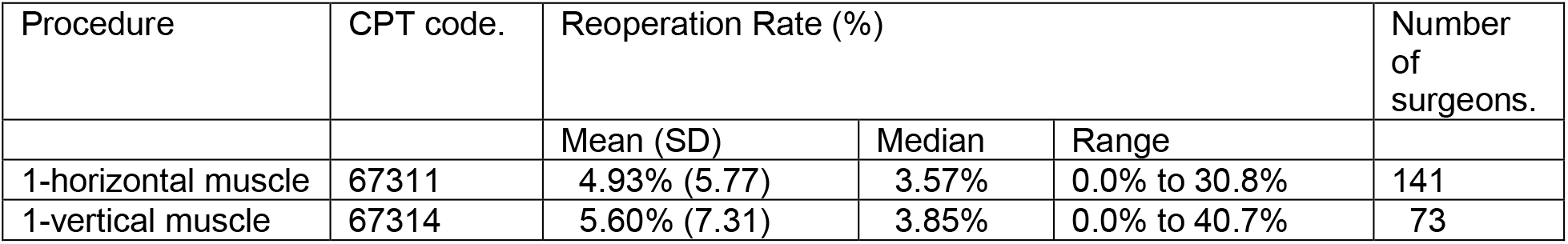
Reoperation rate in the same calendar year for various strabismus procedures.

| Procedure | CPT code. | Reoperation Rate (%) |  |  | Number of surgeons. |
| --- | --- | --- | --- | --- | --- |
|  |  | Mean (SD) | Median | Range |  |
| 1-horizontal muscle | 67311 | 4.93% (5.77) | 3.57% | 0.0% to 30.8% | 141 |
| 1-vertical muscle | 67314 | 5.60% (7.31) | 3.85% | 0.0% to 40.7% | 73 |

<sup>8</sup> Geographic Terms and Concepts-Census Divisions and Census Regions 2017.

**Table 2.** Reoperation rate after one-horizontal muscle surgery (Current Procedural Terminology 67311) by physician.

|  | Reoperation rate, Mean % (SD %, n), when Factor: |  | p value |
| --- | --- | --- | --- |
|  | Present | Absent |  |
| **Surgeon factors:** |  |  |  |
| Peds/strabismus | 4.74% (5.40, 125) | 6.44% (8.21, 16) | 0.43 |
| Neuro-eye | 7.00% (8.65, 14) | 4.71% (5.37, 127) | 0.35 |
| Oculoplastics | 2.53% (1.59, 2) | -- | -- |
| Junior | 3.21% (2.91, 13) | 5.11% (5.97, 128) | 0.06 |
| Mid-career | 5.10% (5.71, 121) | -- | -- |
| Senior | 5.20% (10.09, 7) | 4.92% (5.52, 134) | 0.94 |
| Academic | 4.70% (4.83, 54) | 5.08% (6.31, 87) | 0.69 |
| South | 6.27% (7.30, 60) | 3.94% (4.09, 81) | 0.03 |
| West | 3.68% (4.11, 35) | 5.35% (6.19, 106) | 0.07 |
| Female surgeon | 5.31% (5.68, 34) | 4.88% (5.90, 107) | 0.71 |
| Volume ≥ 37 pts. | 5.14% (4.37, 71) | 4.72% (6.94, 70) | 0.67 |
| Early beneficiaries 2012-16<br>≥50% of total (2012-20) | 5.98% (7.09, 70) | 3.90% (3.87, 71) | 0.03 |
| **Other CPT codes used...** |  |  |  |
| Adjustable suture (67335) | 5.18% (6.54, 46) | 4.82% (5.40, 95) | 0.75 |
| 2-muscle horiz. surgery (67312) | 7.78% (5.47, 18) | 4.52% (5.72, 123) | 0.03 |
| 1-muscle vertical surgery (67314) | 5.59% (5.45, 65) | 4.37% (6.02, 76) | 0.21 |
| Scarring/ reoperation (67332) | 6.05% (4.49, 40) | 4.49% (6.17, 101) | 0.10 |
| **Patient characteristics:** |  |  |  |
| Mean age ≥ 71 years | 5.68% (6.16, 89) | 3.65% (4.85, 52) | 0.03 |
| Female ≥ 57.23% | 5.34% (6.39, 71) | 4.52% (5.09, 70) | 0.40 |
| Medicaid ≥ 15.17% | 3.71% (4.58, 63) | 6.33% (6.71, 63) | 0.01 |
| White race ≥ 86.62% | 6.70% (6.70, 51) | 4.17% (5.41, 50) | 0.04 |
| Diabetes ≥ 25.0% | 5.30% (6.88, 71) | 4.89% (4.43, 60) | 0.68 |
| Stroke ≥ 8.0% | 6.37% (6.63, 41) | 4.46% (6.27, 38) | 0.19 |
| CMS-HCC Risk score ≥ 1.16 | 5.58% (6.40, 71) | 4.28% (5.02, 70) | 0.18 |
| **All practices** | 4.93% (5.77, 141) | -- | -- |

The median values for the patient characteristics for each practice were a volume of 37 patients total from 2012 to 2020; Medicaid qualification (an indicator of poverty) of 15.17%; white race of 86.62%, and fraction with diabetes of 25.0%. The median practice CMS Hierarchical Condition Category (HCC) risk score was 1.1604.

Surgeons in the South had a higher reoperation rate (6.27% vs. 3.94%, p=0.03, Table 2).

Some findings were consistent with strabismus etiology impacting reoperation rate, though none of the findings were statistically significant. Surgeons who also coded for 2 horizontal muscles in 1 eye (CPT 67312) tended to have a higher reoperation rate (7.78% vs. 4.52%, p=0.03, Table 2). Neuro-ophthalmologists tended to have a higher reoperation rate (7.00% vs. 4.71%, p=0.35), as did surgeons with a prevalence of stroke in their practice above the median value (reoperation rate 6.37% vs. 4.46%, p=0.19), but these findings were not statistically significant (Table 2).

Inexperience did not appear to be associated with a higher reoperation rate. The reoperation rate was not higher for low-volume practices (rate 4.72%) below the median value (37 patients from 2012 to 2020), compared to higher-volume practices (rate 5.14%, p=0.67, Table 2, Figure 2). Also, junior surgeons (rate 3.21%) did not have a higher reoperation rate than other surgeons (5.11%, p=0.06 Table 2).

**Figure 2.**
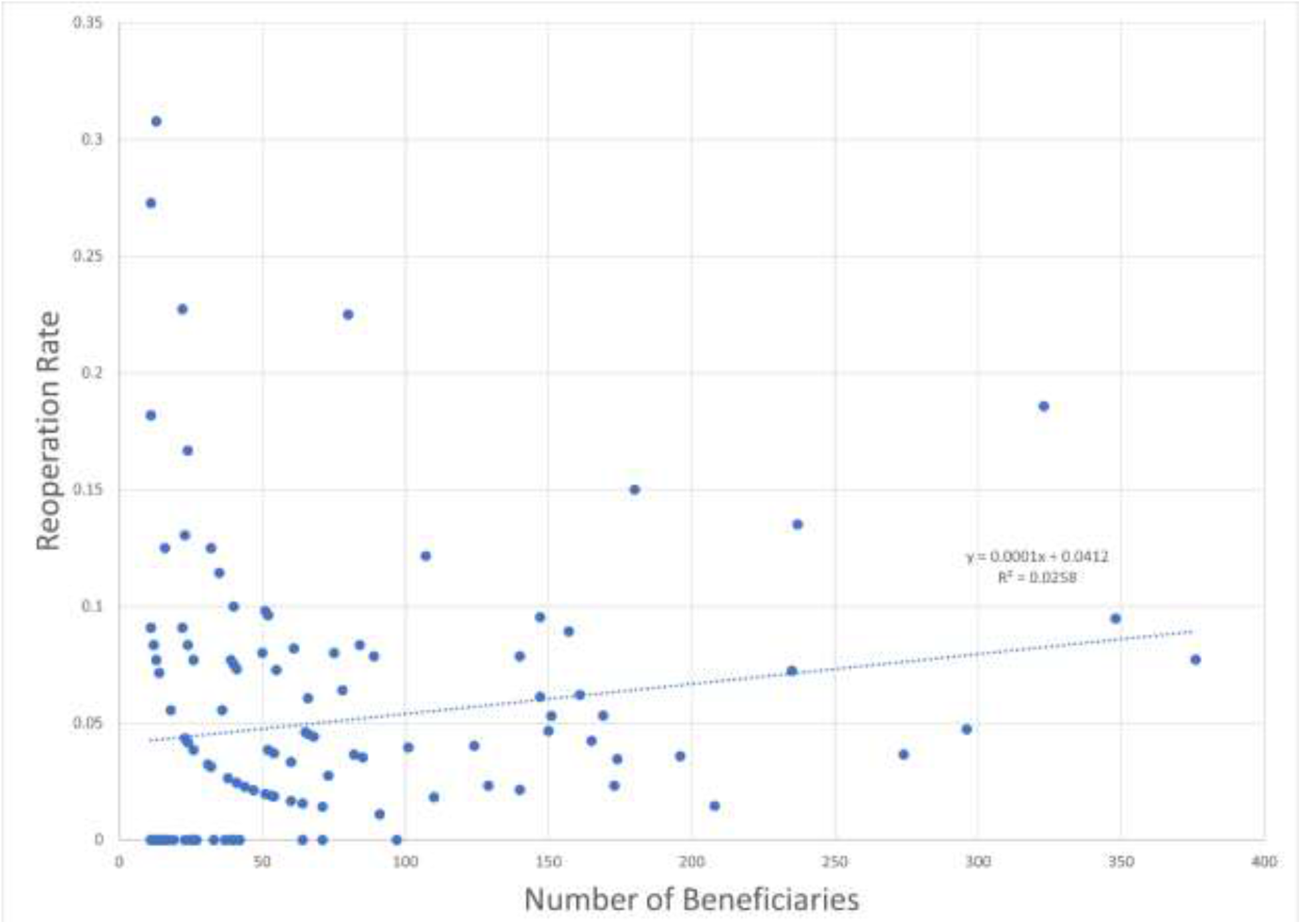
Funnel plot of same calendar year reoperation rate as a function of surgical volume for 1-horizontal muscle surgery (CPT 67311) from 2012 to 2020 for 141 surgeons.

Because of the presence of high-volume, high-reoperation surgeons, a relatively small number of surgeons contributed a substantial fraction of the reoperations in the dataset. For horizontal muscle surgery (CPT 67311), just 11 surgeons of the 141 coding for CPT 67311 (7.8%) contributed 50.4% of the total number of reoperations in the dataset (Figure 3). Three of these 11 high-influence surgeons used adjustable sutures.

**Figure 3.**
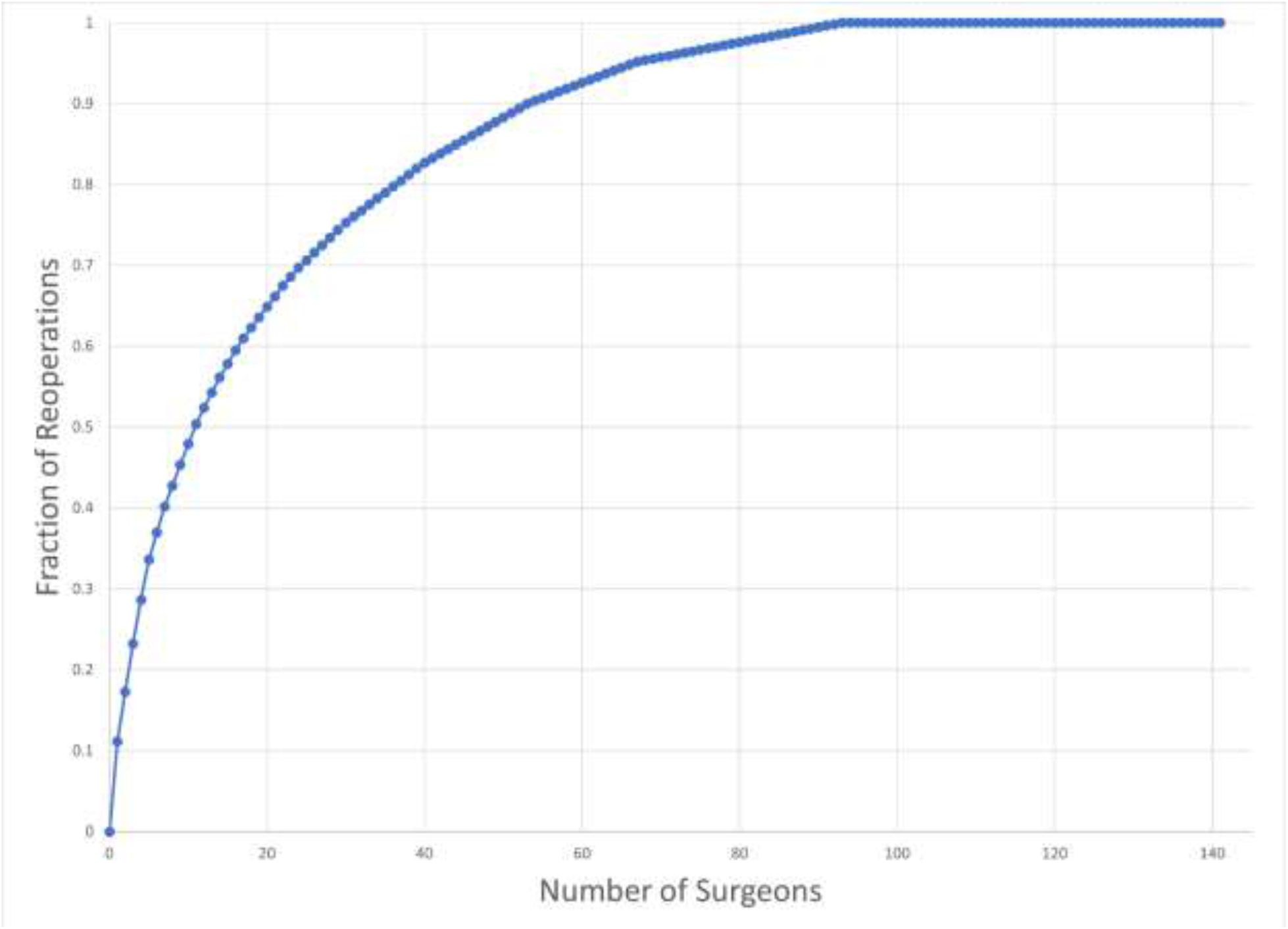
Cumulative contribution of reoperations to the dataset for 141 surgeons for one-horizontal muscle surgery (CPT 67311).

**Figure 4.**
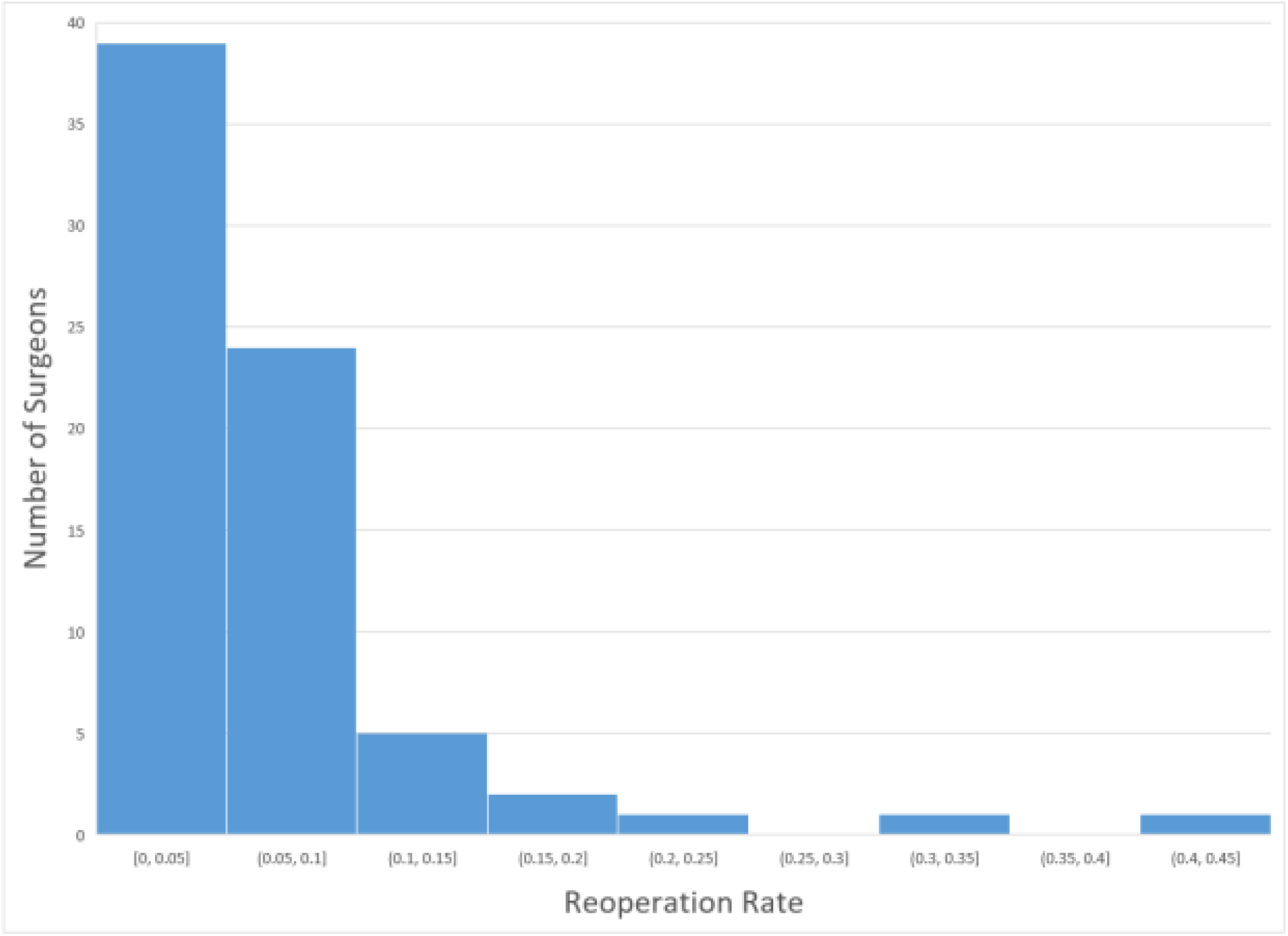
Histogram of reoperation rate among 73 surgeons for one-vertical muscle surgery (CPT 67314).

The 46 surgeons who billed for adjustable sutures (CPT 67335) had a slightly higher reoperation rate than the 95 surgeons who did not (5.18% vs. 4.82%, p=0.75, Table 2). Similarly, when controlling for other practice variables, the availability of adjustable sutures was associated with a nonsignificant elevation in reoperation rate of 0.75% (p=0.57, Table 3).

**Table 3.** Multivariable prediction of reoperation rate (%) after one-horizontal muscle surgery (CPT 67311).

|  | Regression coefficient<br>(95% CI) | p value |
| --- | --- | --- |
| Adjustable sutures used (CPT 67335) | 0.75 (-1.87 to 3.38) | 0.57 |
| Two-horiz. muscle surgery used (CPT 67312) | 1.65 (-1.70 to 5.01) | 0.33 |
| Practice in the South | 2.73 (0.23 to 5.23) | 0.03 |
| Early beneficiaries 2012-16<br>≥50% of total (2012-20) | 1.95 (-0.39 to 4.29) | 0.10 |
| Patient population: |  |  |
| ...Mean age ≥ 71 years. | 1.06 (-2.10 to 4.23) | 0.51 |
| ...Medicaid ≥ 15.17% | -1.26 (-4.38 to 1.86) | 0.43 |
| ...White patients ≥ 86.62% | 1.50 (-1.16 to 4.17) | 0.26 |
| Intercept | 1.74 (-2.53 to 6.02) | 0.42 |

n=101 surgeons analyzed with complete data. Model $r^2=0.163$ .

By multivariable regression, practice in the South was associated with a reoperation rate 2.73% higher (p=0.03), and predominance of beneficiaries from the early years (2012-16) of the total dataset (2012-20) was associated with an elevation in the reoperation rate of 1.96%, which was not statistically significant (p=0.10). The multivariable analysis explained only 16.3% of the inter-surgeon variation in reoperation rate.

### Vertical Muscle Surgery

For vertical muscle surgery, inexperience was not associated with a higher reoperation rate. Higher-volume practices did not have a lower reoperation rate (Table 4, Figure 5). Nor did junior surgeons have a higher reoperation rate (Table 4).

**Table 4.** Reoperation rate after one-vertical muscle surgery (Current Procedural Terminology 67314).

|  | Reoperation rate, Mean % (SD %, n), when Factor: |  | p value |
| --- | --- | --- | --- |
|  | Present | Absent |  |
| **Surgeon factors:** |  |  |  |
| Peds/strabismus | 5.23% (6.16, 62) | 7.63% (12.16, 11) | 0.54 |
| Neuro-eye | 8.39% (12.54, 10) | 5.15% (6.15, 63) | 0.44 |
| Oculoplastics | 0.0% (--, 1) | -- | -- |
| Junior | 1.60% (3.58, 5) | 5.89% (7.44, 68) | 0.053 |
| Mid-career | 5.60% (7.11, 67) | -- | -- |
| Senior | 25.0% (--, 1) | 5.33% (6.99, 72) | -- |
| Academic | 5.81% (7.52, 36) | 5.39% (7.20, 37) | 0.81 |
| South | 5.74% (5.41, 24) | 5.52% (8.13, 49) | 0.89 |
| West | 6.45% (8.03, 24) | 5.17% (6.98, 49) | 0.51 |
| Female surgeon | 5.80% (8.56, 14) | 5.55% (7.06, 59) | 0.92 |
| Volume ≥ 29 pts. | 6.62% (4.46, 37) | 4.54% (9.34, 36) | 0.23 |
| Early beneficiaries<br>2012-16 ≥50% of total<br>(2012-20) | 4.96% (7.94, 38) | 6.29% (6.61, 35) | 0.44 |
| **Other CPT codes**<br>**used...** |  |  |  |
| Adjustable suture (67335) | 5.23% (6.17, 33) | 5.90% (8.20, 40) | 0.69 |
| 2-muscle horiz. surgery<br>(67312) | 7.10% (4.94, 18) | 5.10% (7.91, 55) | 0.21 |
| Scarring/ reoperation<br>(67332) | 7.51% (7.23, 34) | 3.93% (7.05, 39) | 0.04 |
| **Patient characteristics:** |  |  |  |
| Mean age ≥ 72 years | 5.47% (6.45, 37) | 5.72% (8.19, 36) | 0.89 |
| Female ≥ 58.04% | 5.42% (9.30, 36) | 5.77% (4.77, 37) | 0.84 |
| Medicaid ≥ 15.07% | 2.98% (4.05, 27) | 8.01% (9.38, 32) | 0.008 |
| White race ≥ 86.62% | 6.62% (8.97, 27) | 5.83% (7.38, 24) | 0.73 |
| Diabetes ≥ 25.0% | 5.22% (7.26, 27) | 6.52% (7.55, 41) | 0.48 |
| Stroke ≥ 8.0% | 5.81% (4.38, 22) | 4.82% (7.09, 21) | 0.59 |
| CMS-HCC Risk score ≥<br>1.16 | 6.85% (8.58, 33) | 4.44% (5.78, 38) | 0.17 |
| **All practices** | 5.60% (7.31, 73) | -- | -- |

The median values for the patient characteristics for each practice were a volume of 29 patients total from 2012 to 2020.

**Figure 5.**
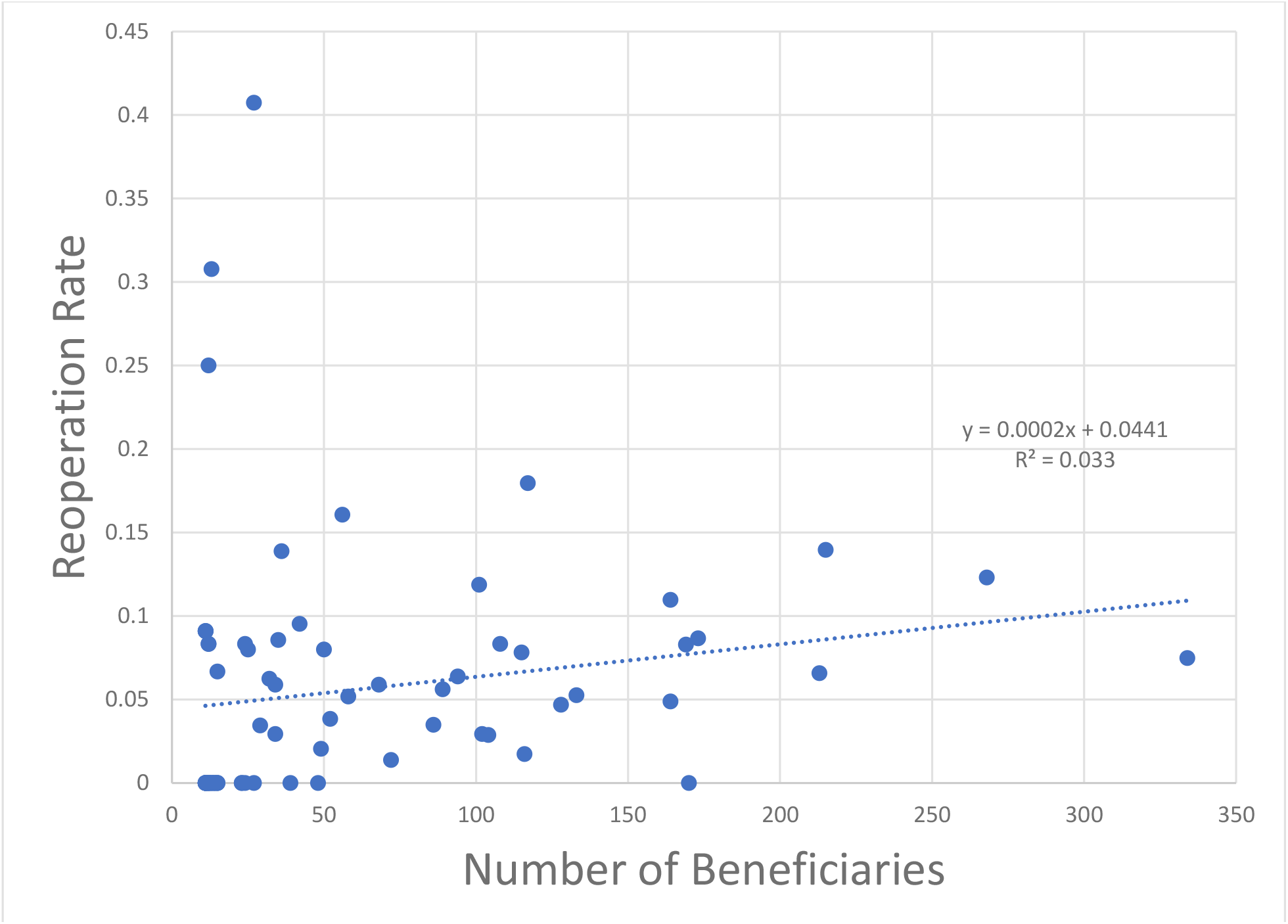
Funnel plot of same calendar year reoperation rate as a function of surgical volume for 1-vertical muscle surgery (CPT 67314) from 2012 to 2020 for 73 surgeons.

**Figure 6.**
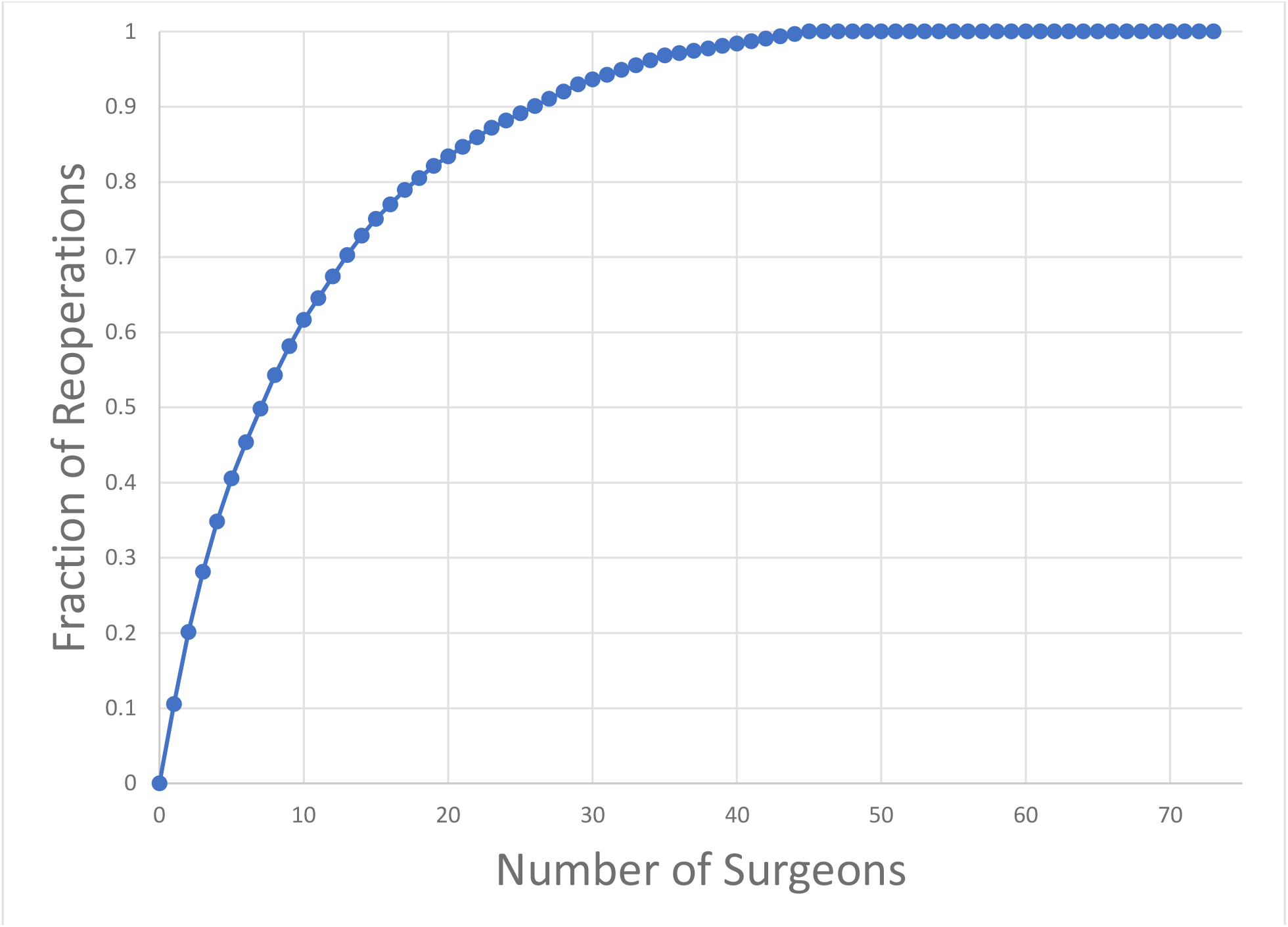
Cumulative contribution of reoperations to the dataset for 73 surgeons for one-vertical muscle surgery (CPT 67314).

For vertical surgery, patient poverty, as evidenced by a practice Medicaid enrollment of at least 15.07%, was associated with a lower reoperation rate of 2.98%, as compared with a rate of 8.01% for comparison practices (p=0.008, Table 4). This association continued to be statistically significant in multivariable analysis (p=0.03, Table 5).

**Table 5.**
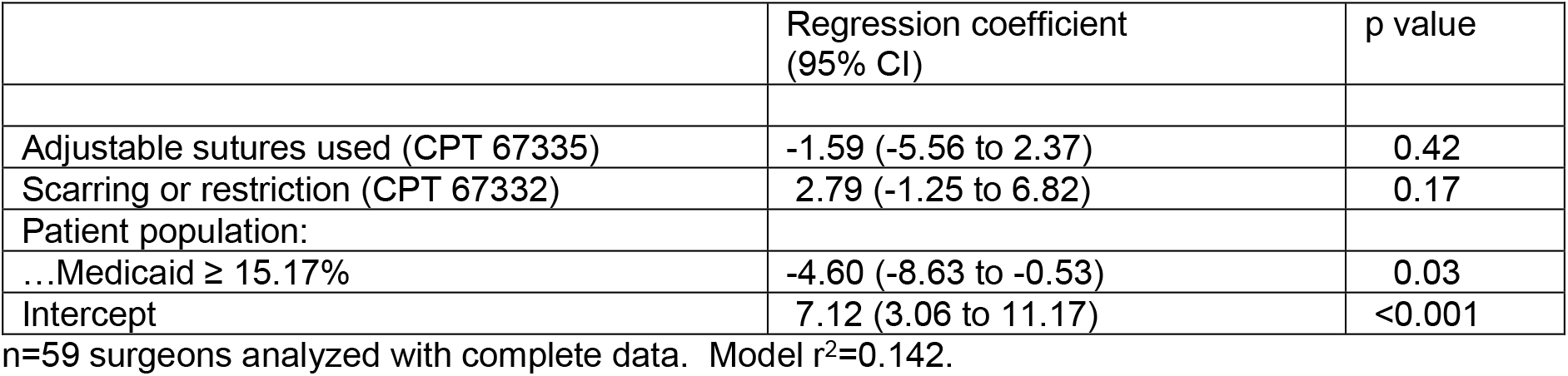
Multivariable prediction of reoperation rate (%) after one-vertical muscle surgery (CPT 67314).

Because of the presence of high-volume, high-reoperation surgeons, a relatively small number of surgeons contributed a substantial fraction of the reoperations in the dataset. For vertical muscle surgery (CPT 67314), a total of 314 reoperations were observed in 4438 cases (7.08%). Just 8 surgeons of the 73 coding for CPT 67314 (10.8%) contributed 54.3% of the total number of reoperations in the dataset (Figure 3). Four of these 8 high-influence surgeons (50%) used adjustable sutures.

## Discussion

This study examined differences between surgeons in reoperation rate following strabismus surgery in older adults.

One of the paradoxical findings is that strabismus surgery reoperation rate does not obey the reductions which would be expected with experience. Reoperation rate was not lower for higher volume surgeons, or for those who were more senior in their career.

A corollary is that high-volume surgeons with high reoperation rates in the dataset tend to dominate any patient-level analyses which ignore inter-surgeon differences. Just 11 surgeons contributed half the reoperation events for 1-horizontal muscle surgery in this national database. Thus, patient-level analyses which ignore the surgeon factor might be describing idiosyncratic practice patterns of a handful of surgeons without yielding generalizable knowledge.

Geographic variation in levels of strabismus surgery,^9^ and outcomes from the procedure, have been demonstrated, though with some inconsistencies.^10^ High-reoperation practices can introduce spurious results when practice variation is not considered. In the present study, each surgeon contributed one observation to the average, and less regional variation was observed, although surgeons in the South did have a higher reoperation rate for horizontal muscle surgery.

Previous studies have found that paralytic strabismus was associated with higher reoperation rates than non-paralytic strabismus.^11^ Likewise, the present study noted higher horizontal surgery reoperation rates in practices which billed for recess-resect procedures (CPT 67312) in univariate analysis. We also noted nonsignificant tendencies for higher reoperation rates among neuro-ophthalmologists and practices with a higher prevalence of stroke.

Older patient age was associated with a higher reoperation rate in our study and previous studies.^12^ We found that patient poverty was associated with a lower reoperation rate for vertical muscle surgery. We are unaware of previous studies examining this association.

Reoperation rates vary between surgeons, between zero and 30.8% in the first calendar year for horizontal surgery, and between zero and 40.7% for vertical surgery. Despite the many patient and surgeon variables explored, most of the variation in this outcome between surgeons remains unexplained.

## Data Availability

Data are available from the first author.

## Footnotes

1 Schrag 2002, Cowan 2003.

2 Bell 2007, Johnston 2010.

3 Hopkinson 2022.

4 Leffler “Digit J” 2016; Christensen, Pierson, Leffler 2018; Leffler AJO 2015; Leffler AJO 2016; Repka 2018; Oke 2022; Colas 2022.

5 Medicare Provider Utilization and Payment Data 2022.

6 Medicare Provider Utilization and Payment Data 2022.

7 Medicare Physician & Other Practitioners - by Provider 2022.

8 Geographic Terms and Concepts-Census Divisions and Census Regions 2017.

9 Chou 2013, Repka 2013.

10 Christensen, Pierson, and Leffler 2018; Repka 2018; Oke 2022.

11 Leffler AJO 2015; Colas 2022.

12 Leffler AJO 2015; Oke 2022.

